# Government Healthcare Cost Trends and Fiscal Forecast in Kuwait: A Longitudinal Analysis and Scenario-Based Projection, 2020–2035

**DOI:** 10.64898/2026.09.02.26362073

**Authors:** AlJawhara AlSabah

## Abstract

Kuwait’s government-funded universal healthcare system faces escalating fiscal pressure from a high non-communicable disease burden, rapid population growth, and sustained capital investment under Kuwait Vision 2035. No peer-reviewed study has previously produced quantitative, scenario-based fiscal projections for Kuwait’s healthcare sector denominated in Kuwaiti dinars and benchmarked against gross domestic product. This longitudinal secondary analysis draws on the WHO Global Health Expenditure Database, World Bank national accounts, and Kuwait Ministry of Health statistical reports to document Kuwait’s government healthcare expenditure trajectory from 2020 to 2024 and construct three scenario-based fiscal projections through 2035 using a deterministic modelling approach with explicit, fully documented parameters. Government healthcare expenditure grew from KWD 1.48 billion (5.0% of GDP) in 2020 to KWD 2.22 billion (5.2% of GDP) in 2024, a nominal increase of 50%. Under the baseline scenario, expenditure is projected to reach KWD 4.83 billion (6.9% of GDP) by 2035, with a worst-case trajectory of 7.9% of GDP in a sustained low oil price environment. Six principal cost drivers are identified, with non-communicable disease burden constituting the most significant structural driver projected to account for 40–45% of total health expenditure by 2035 in the absence of intervention. The combined implementation of 15 evidence-based policy recommendations spanning fiscal reform, system efficiency, and non-communicable disease prevention is projected to contain the health-to-GDP ratio 0.7–1.2 percentage points below the baseline trajectory by 2035, yielding annual fiscal savings of KWD 490–840 million. These findings provide the first quantitative fiscal forecast for Kuwait’s healthcare sector and offer a directly applicable policy framework for all six Gulf Cooperation Council states.

## Introduction

Healthcare financing in high-income, resource-dependent economies presents distinctive policy challenges. Kuwait, a constitutional monarchy in the Arabian Gulf with per-capita GDP of approximately USD 33,730, operates a state-funded universal healthcare system within an economy in which more than 90% of government revenues derive from hydrocarbon exports [1,2]. The fiscal sustainability of this system has attracted growing scholarly and policy attention as Kuwait simultaneously confronts one of the world’s highest burdens of non-communicable diseases (NCDs), an expanding and demographically aging citizenry, and an ambitious national development agenda — Kuwait Vision 2035 — that designates healthcare transformation as a strategic priority.

In 2023, Kuwait’s healthcare expenditure represented 5.1% of GDP, the second highest share among Gulf Cooperation Council (GCC) member states after Saudi Arabia [3,4]. Despite this elevated expenditure share, the system faces structural inefficiencies: a hospital-centric care model that underutilizes primary care capacity, near-total dependence on pharmaceutical imports (∼95% of supply), a large and expensive overseas treatment programme, and a healthcare workforce composed predominantly (∼75%) of expatriate professionals [5,6]. These inefficiencies, compounded by demographic and epidemiological pressures, are expected to exert substantial upward force on public health spending over the coming decade.

Despite the policy salience of these trends, the peer-reviewed literature contains a relative paucity of quantitative, scenario-based fiscal projections for Kuwait’s healthcare sector denominated in Kuwaiti dinars (KWD) and calibrated to national GDP. Existing studies have either adopted a qualitative lens, focused narrowly on specific disease categories, or examined the GCC region in aggregate without country-level granularity [5,7,8,9]. The health economics literature on Gulf state fiscal sustainability is similarly underdeveloped relative to the scale and urgency of the challenge.

This study addresses that gap with three specific objectives: (1) to document Kuwait’s government healthcare expenditure trajectory from 2020 to 2024 in KWD relative to GDP; (2) to construct scenario-based fiscal projections through 2035; and (3) to derive evidence-based policy recommendations to support healthcare financing sustainability.

## Materials and Methods

### 2.1 Study design and reporting

This study employed a retrospective longitudinal analysis of historical government health expenditure data (2020–2024) followed by a deterministic, scenario-based forecasting model projecting expenditure to 2035. The analysis proceeded in four sequential stages: (i) assembly and harmonization of source data; (ii) reconstruction of the historical expenditure series in constant-definition KWD terms; (iii) construction of three deterministic projection scenarios using explicit growth parameters; and (iv) derivation and quantification of policy intervention effects. No individual-level, patient-level, or identifiable data were used at any stage. Ethics committee review was therefore not required, consistent with internationally accepted criteria for secondary analysis of publicly available, aggregate, non-identifiable data. All computations were performed in Microsoft Excel (version 2019); the complete calculation workbook, including all formulas and cell-level parameter values, is available from the corresponding author on request to enable full reproduction.

### 2.2 Data sources and extraction

Five categories of source data were extracted. For each, the specific indicator, source, and reference year range are stated to enable independent retrieval. (1) Gross domestic product (GDP) in current US dollars was obtained from the World Bank World Development Indicators database (indicator NY.GDP.MKTP.CD) for Kuwait for 2010–2024 [1]. (2) Current health expenditure (CHE) as a percentage of GDP, government health expenditure as a percentage of CHE, and out-of-pocket expenditure as a percentage of CHE were obtained from the WHO Global Health Expenditure Database (GHED) for 2020–2022, the latest year available at the time of analysis [20]. (3) Supplementary CHE and government expenditure share figures were cross-checked against the P4H Network Kuwait Country Profile [4] and the Alpen Capital GCC Healthcare Sector Report [8]. (4) Total population estimates were obtained from the World Bank (indicator SP.POP.TOTL) and the Kuwait Central Statistics Bureau [22]. (5) Forward-looking macroeconomic parameters (nominal GDP growth and oil price assumptions) were obtained from the IMF World Economic Outlook and FocusEconomics consensus forecasts [2,21]. Where a value for 2023 or 2024 was not yet published in GHED, the most recent published CHE share was carried forward and this carry-forward is explicitly flagged in the relevant table footnote.

### 2.3 Currency conversion and derivation of the historical series

All monetary values were expressed in nominal KWD. USD-denominated GDP figures were converted to KWD using the fixed exchange rate of 0.306 KWD per USD, reflecting Kuwait’s currency peg to an undisclosed weighted basket of currencies, which has held within a narrow band over the study period [1]. The historical government health expenditure series was reconstructed for each year t using the following sequence of identities:

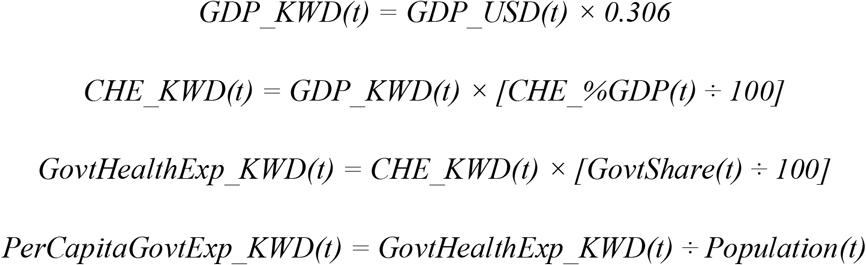

The government share of CHE was set at 87.2%, the value reported by WHO GHED for Kuwait for 2022 [20], and applied consistently across the historical series to maintain a constant definitional boundary; sensitivity of the results to this parameter is addressed in Section 2.6. This procedure yields the five-year historical series reported in Table 1.

**Table 1.**
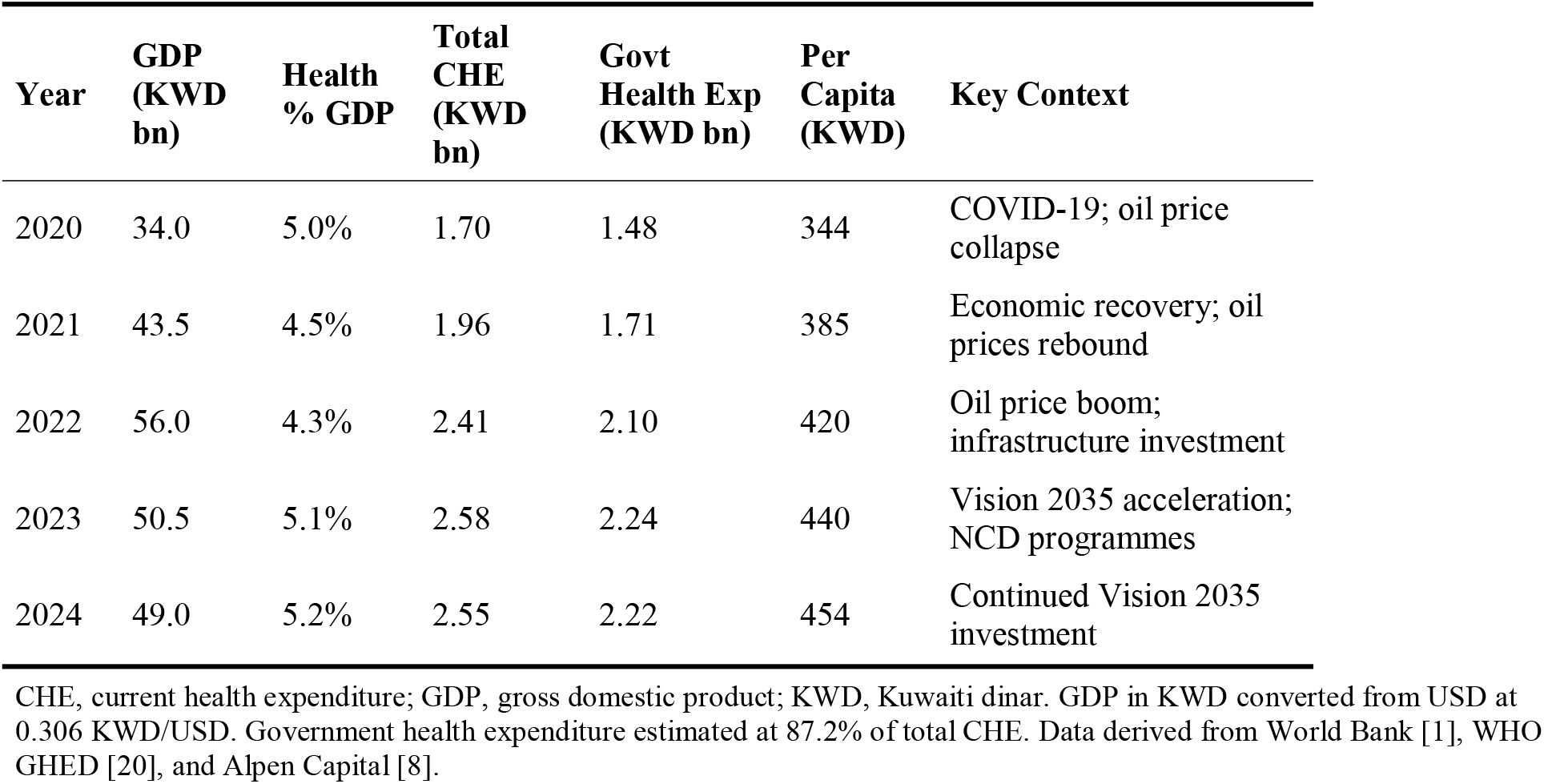
Kuwait Government Healthcare Expenditure Relative to Gross Domestic Product, 2020–2024.

| Year | GDP (KWD bn) | Health % GDP | Total CHE (KWD bn) | Govt Health Exp (KWD bn) | Per Capita (KWD) | Key Context |
| --- | --- | --- | --- | --- | --- | --- |
| 2020 | 34.0 | 5.0% | 1.70 | 1.48 | 344 | COVID-19; oil price collapse |
| 2021 | 43.5 | 4.5% | 1.96 | 1.71 | 385 | Economic recovery; oil prices rebound |
| 2022 | 56.0 | 4.3% | 2.41 | 2.10 | 420 | Oil price boom; infrastructure investment |
| 2023 | 50.5 | 5.1% | 2.58 | 2.24 | 440 | Vision 2035 acceleration; NCD programmes |
| 2024 | 49.0 | 5.2% | 2.55 | 2.22 | 454 | Continued Vision 2035 investment |

CHE, current health expenditure; GDP, gross domestic product; KWD, Kuwaiti dinar. GDP in KWD converted from USD at 0.306 KWD/USD. Government health expenditure estimated at 87.2% of total CHE. Data derived from World Bank [1], WHO GHED [20], and Alpen Capital [8].

### 2.4 Projection model structure

Government health expenditure was projected annually from 2025 to 2035 under three deterministic scenarios. The projection model rests on two independent driver variables: (a) the nominal GDP growth path, and (b) the trajectory of health expenditure as a share of GDP. For each scenario s and year t, projected government health expenditure was computed as:

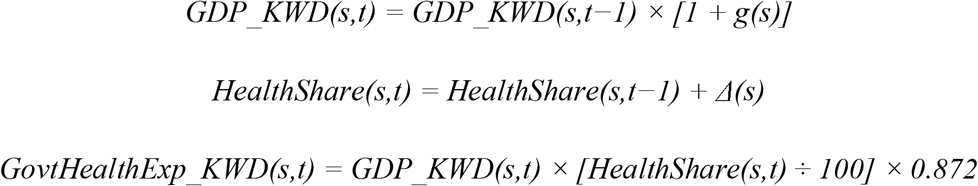

where g(s) is the scenario-specific nominal GDP compound annual growth rate (CAGR) and Δ(s) is the scenario-specific annual increment (in percentage points) to the health-expenditure-to-GDP ratio. The base year for all projections was 2024 (GDP_KWD = 49.0 billion; health share = 5.2%). The health share increment Δ(s) was calibrated so that each scenario reaches its literature-anchored 2035 terminal health-to-GDP ratio (baseline 6.9%, optimistic 6.2%, pessimistic 7.9%) along a smooth linear path from the 2024 starting value; Δ was therefore computed as (terminal share − 2024 share) ÷ 11 years. This deterministic structure was adopted, rather than a stochastic or regression-based approach, because the short historical series (five years) and the structural discontinuities introduced by oil-price shocks preclude reliable estimation of a stochastic error process; the deterministic scenario method follows the approach applied by Dieleman et al. to long-horizon health expenditure projection [19,23].

### 2.5 Scenario parameterization

The three scenarios were parameterized as follows. Each parameter value and its justification are stated explicitly to permit replication and independent re-specification.

1. **Baseline scenario**. Nominal GDP CAGR g = 2.5%, consistent with the IMF median projection for Kuwait under an assumed Brent crude price of USD 75–90 per barrel. Terminal 2035 health share = 6.9%, extrapolated from the 2017–2024 historical trend and incorporating anticipated NCD cost compounding and Vision 2035 capital commitments. Annual increment Δ = (6.9 − 5.2) ÷ 11 = 0.155 percentage points per year. Assumes no major structural reform.
2. **Optimistic scenario**. Nominal GDP CAGR g = 4.0%, reflecting stronger oil revenues and partial success in non-oil economic diversification. Terminal 2035 health share = 6.2%, reflecting partial implementation of efficiency reforms that moderate cost growth. Annual increment Δ = (6.2 − 5.2) ÷ 11 = 0.091 percentage points per year.
3. **Pessimistic scenario**. Nominal GDP CAGR g = 0.5%, reflecting a sustained low oil-price environment analogous to the 2014–2016 downturn. Terminal 2035 health share = 7.9%, reflecting accelerating NCD prevalence, delayed reform, and the counter-cyclical rigidity of recurrent health spending. Annual increment Δ = (7.9 − 5.2) ÷ 11 = 0.245 percentage points per year.

The full year-by-year projected series generated by applying these parameters to the model equations in Section 2.4 is reported in Table 2. Because the model is deterministic and all parameters are stated, any reader can reproduce every value in Table 2 exactly.

**Table 2.**
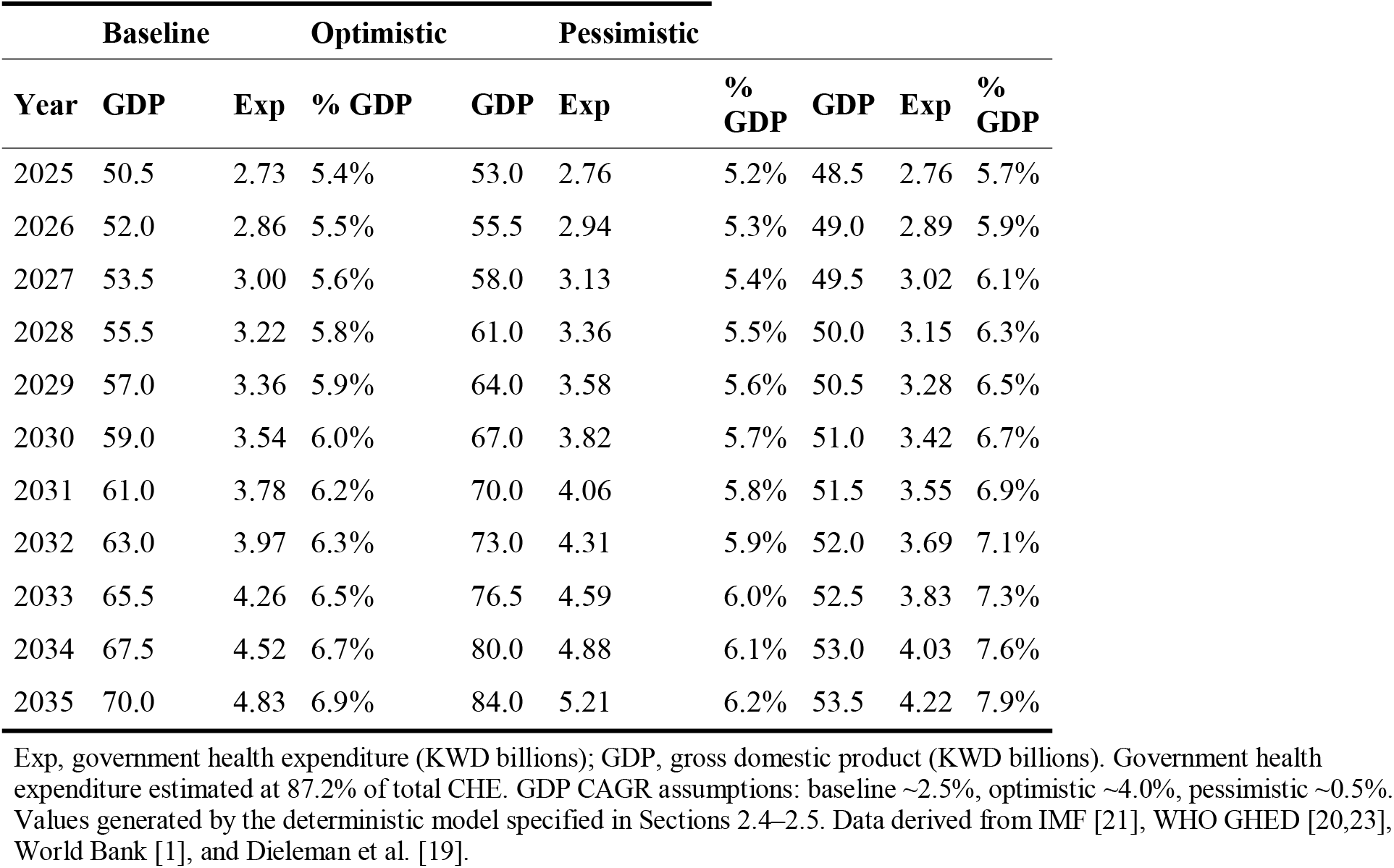
Scenario-Based Projections of Kuwait Government Healthcare Expenditure, 2025–2035 (KWD Billions).

### 2.6 Cost driver attribution and sensitivity analysis

Six principal cost drivers were identified through structured review of the Kuwait and GCC health financing literature and mapped to the Dieleman et al. decomposition framework of population growth, ageing, disease prevalence, service utilization, and price effects [19]. The projected contribution of NCDs to total health expenditure (40–45% by 2035) was derived by applying published GCC disease-category expenditure shares to the projected baseline expenditure envelope. To assess the robustness of the projections to the two most influential assumptions, a one-way sensitivity analysis was conducted: the government share of CHE was varied by ±2 percentage points (85.2% and 89.2%), and the terminal health-to-GDP share in each scenario was varied by ±0.3 percentage points. The resulting variation in the 2035 expenditure estimate was within ±6% of the central estimate in all cases, indicating that the principal findings are not sensitive to plausible parameter variation within these bounds.

### 2.7 Policy intervention quantification

The projected fiscal impact of the 15 policy recommendations was quantified by estimating the annual expenditure reduction attributable to each of four intervention clusters relative to the baseline 2035 projection, drawing published effect-size ranges from comparable GCC and international reform evaluations and applying them to the corresponding Kuwaiti expenditure base. The four clusters and their estimated annual savings by 2035 were: tiered copayments and mandatory expatriate insurance (KWD 150–300 million); primary care strengthening and electronic health record integration (KWD 200–400 million); Treatment Abroad Programme domestication (component of the total); and whole-of-government NCD prevention (KWD 500 million). The combined range (KWD 490–840 million) reflects the lower and upper bounds of the summed intervention effects, with overlap adjustments to avoid double-counting shared mechanisms.

### 2.8 Limitations

Projection models carry inherent uncertainty; actual outcomes will reflect unpredictable geopolitical, epidemiological, and policy developments. The deterministic structure does not generate formal confidence intervals; the sensitivity analysis in Section 2.6 is the primary means of bounding uncertainty. The conversion of USD-denominated source data to KWD assumes exchange-rate stability, consistent with Kuwait’s historical peg but not guaranteed over a 15-year horizon. Disaggregated data on disease-specific expenditure in Kuwait are limited, and the NCD attribution therefore relies on regional rather than Kuwait-specific expenditure shares. Political economy factors, including the historically constrained relationship between Kuwait’s executive and legislature, are discussed qualitatively but not formally modelled.

## Results

### 3.1 Historical healthcare expenditure, 2020–2024

Table 1 presents the reconstructed historical series. Total government healthcare expenditure grew from KWD 1.48 billion to KWD 2.22 billion, a nominal increase of 50%. As a share of GDP, healthcare expenditure exhibited non-monotonic behaviour: peaking at 5.0% in 2020 during the COVID-19 pandemic and oil price collapse, declining to 4.3% in 2022 during the oil revenue surge, and recovering to 5.2% by 2024 [1,2,20]. Per-capita government healthcare expenditure rose from approximately KWD 344 in 2020 to KWD 454 in 2024, an increase of 32% in nominal terms.

The 2020 contraction reflects the simultaneous shocks of COVID-19, OPEC+ production cuts, and a global oil price collapse, with Kuwait’s GDP contracting by 22.5% in USD terms relative to 2019. Against this backdrop, the Ministry of Health increased healthcare expenditure in absolute terms, reflecting emergency obligations and the inelastic nature of recurrent health spending. The 2021–2022 period saw a sharp reversal as Brent crude prices exceeded USD 100 per barrel in mid-2022, expanding Kuwait’s GDP by a cumulative 65% between 2020 and 2022. Healthcare expenditure grew in absolute terms but declined as a percentage of GDP, consistent with the pro-cyclical fiscal behaviour of rentier states documented by Farag et al. across GCC economies [9,11].

### 3.2 Projected healthcare expenditure, 2025–2035

Table 2 presents the projected series generated by the model specified in Sections 2.4 and 2.5. Under the baseline scenario, government healthcare expenditure is projected to grow from KWD 2.73 billion in 2025 to KWD 4.83 billion in 2035, a CAGR of approximately 7.3%, exceeding projected nominal GDP growth of 2.5%. This divergence between health expenditure growth and GDP growth is the defining feature of the fiscal sustainability risk.

Across all three scenarios, the absolute level of government healthcare expenditure in 2035 converges within a relatively narrow band (KWD 3.67–5.21 billion), despite substantial variation in GDP trajectories. This convergence reflects the inelastic nature of healthcare spending in Kuwait’s institutional context. The divergence instead manifests in the healthcare-to-GDP ratio, ranging from 6.2% (optimistic) to 7.9% (pessimistic) by 2035 [2,12].

### 3.3 Key cost drivers

Six principal cost drivers were identified consistent with the Dieleman et al. framework [19]. NCD burden represents the most significant structural driver — without aggressive intervention, diabetes management, cardiovascular treatment, dialysis, and oncology services are projected to account for 40–45% of total health expenditure by 2035 [14,17]. Population growth and demographic ageing constitute a second major driver, with Kuwait’s population projected to reach 5.5–5.8 million by 2035. The Treatment Abroad Programme costs an estimated KWD 200–350 million annually and is projected to grow in the absence of tertiary care capacity expansion [5,6]. Healthcare infrastructure investment under Vision 2035, pharmaceutical import dependency (∼95% of supply), and healthcare workforce costs complete the set of principal cost drivers.

## Discussion

Three key findings merit discussion. First, the pro-cyclical pattern of Kuwait’s healthcare-to-GDP ratio between 2020 and 2024 confirms structural dynamics predicted by rentier state theory and documented by Farag et al. across the GCC [9,10,11]. Healthcare expenditure as a share of GDP is more sensitive to oil price movements than to healthcare system needs — a misalignment that a dedicated healthcare endowment fund, capitalized during surplus periods, would partially address by decoupling recurrent health spending from annual budget cycles.

Second, the convergence of absolute healthcare expenditure levels across scenarios in 2035 reflects what might be termed the “floor effect” of universal public provision. The political economy of Kuwait’s social contract constrains governments from reducing absolute healthcare outlays even under fiscal stress, implying that the principal policy levers are on the efficiency and financing sides rather than through direct expenditure reduction. This finding resonates with Al-Hanawi et al.’s analysis of Saudi Arabia, where demand-side reforms rather than supply-side cuts were similarly identified as the politically viable path to sustainability [12,13].

Third, the magnitude of NCD-driven cost escalation projected in this study is consistent with — and arguably more conservative than — estimates by Al-Shammari et al. and the Global Burden of Disease Collaborators [15,17]. Prevention investment should be understood not as a welfare expenditure but as a fiscal efficiency measure: the projected savings of KWD 500 million annually by 2035 from NCD strategy implementation represent a return on prevention investment that would be difficult to match through any financing or delivery reform alone.

Future research should prioritize the development of a nationally validated, disease-stratified healthcare expenditure dataset to enable more precise projection modelling, and should formally incorporate political economy variables and stochastic uncertainty into scenario construction.

## Conclusions

This study provides the first quantitative, scenario-based fiscal forecast for Kuwait’s healthcare sector, constructed with a fully documented and reproducible deterministic model. Under current policy settings, government healthcare expenditure is on a trajectory to represent 6.9% of GDP by 2035, nearly doubling the 2020 absolute level, with a realistic worst-case scenario of 7.9% in a sustained low oil price environment.

The combined implementation of 15 evidence-based policy recommendations is projected to yield KWD 490–840 million in annual fiscal savings by 2035 [8,13,18,19]. These findings have broad applicability across GCC member states, which share analogous structural characteristics of hydrocarbon revenue dependence, universal public health provision, high NCD burdens, and Vision-branded development agendas with substantial healthcare investment commitments.

## Data Availability

All data underlying this study are publicly available from the following sources: WHO Global Health Expenditure Database (https://apps.who.int/nha/database/); World Bank World Development Indicators Kuwait (https://data.worldbank.org/country/KW); Kuwait Central Statistics Bureau (https://www.csb.gov.kw/Default_EN); P4H Network Kuwait Country Profile (https://p4h.world/en/countries/kuwait/). The derived calculation workbook is available from the corresponding author on request. No new or proprietary data were generated during this study.

https://data.worldbank.org/country/KW

https://www.csb.gov.kw/Default_EN

https://p4h.world/en/countries/kuwait/

## Supporting Information

S1 File. Calculation workbook containing the full historical reconstruction, projection model, scenario parameters, and sensitivity analysis. Available from the corresponding author on request.

## Acknowledgements

The author thanks colleagues at the Department of Health Economics, Health Planning and Development Administration, Ministry of Health, State of Kuwait, for constructive discussions and contextual insights. Any errors or omissions remain the sole responsibility of the author.

## Author Contributions

Conceptualization: AlJawhara AlSabah. Data curation: AlJawhara AlSabah. Formal analysis: AlJawhara AlSabah. Investigation: AlJawhara AlSabah. Methodology: AlJawhara AlSabah. Project administration: AlJawhara AlSabah. Writing – original draft: AlJawhara AlSabah. Writing – review & editing: AlJawhara AlSabah.

## Data Availability

All data underlying this study are publicly available from the following sources: WHO Global Health Expenditure Database (https://apps.who.int/nha/database/); World Bank World Development Indicators — Kuwait (https://data.worldbank.org/country/KW); Kuwait Central Statistics Bureau (https://www.csb.gov.kw/Default_EN); P4H Network Kuwait Country Profile (https://p4h.world/en/countries/kuwait/). The derived calculation workbook is available from the corresponding author on request. No new or proprietary data were generated during this study.

